# Evolution of Pandemic Cholera at its Global Source

**DOI:** 10.1101/2025.02.03.25321585

**Authors:** Amber Barton, Mokibul Hassan Afrad, Alyce Taylor-Brown, Nisha Singh, Chetan Thakur, Taufiqul Islam, Sadia Isfat Ara Rahman, Marjahan Akhtar, Yasmin Ara Begum, Taufiqur Rahman Bhuiyan, Ashraful Islam Khan, Neelam Taneja, Nicholas R. Thomson, Firdausi Qadri

## Abstract

The seventh pandemic of cholera, caused by the 7^th^ Pandemic El Tor Lineage (7PET) *Vibrio cholerae*, was previously shown to have emanated in three global waves from the Bay of Bengal, bordering Bangladesh and India. However, the respective roles of the Ganges delta and basin regions in seeding these global pandemic waves were not known. We find that while there are transmission events between Bangladesh and India, *V. cholerae* within the two countries has largely evolved separately over the past 20 years, contained by national borders rather than following hydrological features such as the Ganges delta and basin. Evolution within Bangladesh was distinct from that seen in India, involving rapid gain and loss of genes and mobile genetic elements, particularly those involved in phage defence. The loss/gain of these anti-phage elements mirrored loss/gain of anti-defence systems in lytic phage ICP1. Importantly, the loss of these systems was associated with increased risk of severe disease and transmission outside of Bangladesh. Here we show that the Ganges basin, falling across Bangladesh and Northern India, rather than the Ganges delta, acts as a global launch pad for pandemic disease. This completely shifts our understanding of Bangladesh as the purported global source of cholera, and the role of phage in controlling spread of lineages within the current seventh pandemic.

## Introduction

Cholera, an acute diarrhoeal infection caused by *Vibrio cholerae*, has caused seven global pandemics, of which the first six were caused by the classical biotype and thought to disseminate globally from the Ganges delta^1^. At the beginning of the last century the El Tor biotype, named after the El Tor quarantine station in Egypt where it was first observed, began to replace the classical biotype in epidemic disease. The seventh cholera pandemic, officially declared in Indonesia in 1960, is the longest-running pandemic for any pathogen^2^ with the largest recorded epidemics occurring in recent times, including Haiti in 2010^3^ and Yemen in 2016^4^ and 2019^5^. Since 2022 there has been a recent uptick in outbreaks: countries such as Lebanon^6^ and Syria^7^ have reported cholera for the first time; there have been unseasonal outbreaks in Malawi^8^, and Bangladesh saw its largest outbreak ever in 2022^9^.

The seventh pandemic can be attributed to a single discrete genetic lineage named seventh pandemic El Tor lineage (7PET)^10^. While 7PET phenotypically shares the same serogroup as the classical biotype (O1), it emerged independently from a non-epidemic El Tor biotype ancestor following acquisition of the CTXφ phage and two pathogenicity islands (VSP-I and VSP-II)^11.^ Since 1960 7PET has disseminated globally in three overlapping waves originating from the Ganges basin^12^.

Global dissemination of 7PET appears as repeated clonal expansions, with 11 observed introduction events into Africa (termed T1 and T3-T12)^13^, three into Latin America (LAT1-LAT3)^14^ and eight into Europe (EUR1-EUR8)^15^. Whilst the dynamics of *Vibrio cholerae* in specific outbreaks or restricted geographic sites have been explored^5,6,16^, the overall evolutionary dynamic has not been studied longitudinally across the “global home for cholera” in the Ganges basin, spanning India and Bangladesh. What is known is that two 7PET lineages have been consistently observed in India and Bangladesh over the past 15 years, previously named BD1 and BD2^9,17–21^. While their genomes differ by fewer than 150 single nucleotide polymorphisms (SNPs), they show marked variation in mobile genetic elements (MGEs), conferring differences in phage defences, antibiotic resistance and virulence. These include genomic islands such as CTXφ, VSP-II and the superintegron^22^; SXT-ICEs such as ICE^TET^ and ICE^GEN23^, plasmids such as pCNRVC190243^5^, and phage-inducible chromosomal island-like elements (PLEs)^24^.

We collected and sequenced isolates from across Bangladesh (n = 1516, 2014-2023) and North India (n = 794, 2002-2023), to generate the most comprehensive longitudinal dataset of cholera in the Ganges basin to date. For many years, the primary global source of cholera was considered to be the brackish waters of the Ganges Delta and the Bay of Bengal. However, by tracking *Vibrio cholerae* in the Ganges basin we observed that cholera transmission is constrained by national boundaries, and therefore likely population mobility, rather than reflecting the patterns we would expect if clinical disease was linked to a primarily environmental transmission path. We infer that despite high prevalence in Bangladesh, global dissemination of cholera is likely mediated by exportation from India. Within Bangladesh, *V. cholerae* shows unique patterns of evolution, particularly the rapid loss/gain of anti-phage elements which are linked to increased risk of rice water stool or severe dehydration. Furthermore, transmission of lineages outside of Bangladesh to other countries may be compromised by the presence of these genetic elements.

## Results

### *Vibrio cholerae* evolution in Bangladesh is characterised by rapid gain and loss of genes and mobile genetic elements

We sequenced isolates collected during a 2014-2018 nationwide systematic cholera surveillance study from sites across Bangladesh (n=1453) and those collected during an all enteric disease screening campaign at icddr,b Hospital in Dhaka (n=63) where every 50^th^ patient visiting was enrolled and tested for enteric pathogens, resulting in 1477 high-quality genomes. From India we sequenced 791 genomes spanning eight Northern Indian states collected through referral, clinical and surveillance services provided by The Post Graduate Institute of Medical Education and Research (PGIMER; Supplementary document 1; Figure S1). Once placed in the context of 3112 published global genomes, hierarchical clustering using hierBAPS revealed eight BAPS clusters. Five clusters corresponded to previously described lineages LAT1^25^, O139^26^, T9^16^, T10 and BD2^17^ (Figure S2). One BAPS cluster corresponded to wave 1 transmission lineages T1-5 (we denote “W1”) and another to wave 2 transmission lineages T6-8 (denoted “W2”). With more data from the Ganges basin and delta, it is clear that the previously named “BD1” encompasses not only the previously defined BD1 lineage, but also transmission lineages T11-13 and LAT3. We refer to this lineage as supra-BD1 (sBD1). To provide the resolution needed to track *V. cholerae* across the Ganges basin and delta, we sub-divided sBD1 and BD2 into discrete sub-lineages based on hierBAPS clusters at lower levels (e.g. sBD1.1.1).

From the two Bangladesh surveillance studies, BD2 predominated in Bangladesh from 2014-2017 (Figure 1a) before being replaced in 2018 by sBD1, inferred by ancestral reconstruction to have been re-introduced from India (Figure S2). Notably, this is the only major introduction event observed for Bangladesh in this study: all other predominant sub-lineages circulating in Bangladesh were the same as or descended from sub-lineages from the previous year. The persistence of each sub-lineage varied (Figure 1a), with an average duration of 19 months (interquartile range 13-24 months) within the 2014-2018 systematic surveillance study. Of the 16 sub-lineages observed in Bangladesh during the surveillance study, 15 were detected in more than one administrative division. Importantly, considering pathogen migration rates, there was a lag of 14 months (linear model, 95% confidence intervals 12-17 months) between first detection and a sub-lineage spreading to all eight divisions in Bangladesh (Figure S3). Of note, 10/16 sub-lineages were first detected in Chittagong, the southernmost division in Bangladesh bordering the Bay of Bengal.

**Figure 1:**
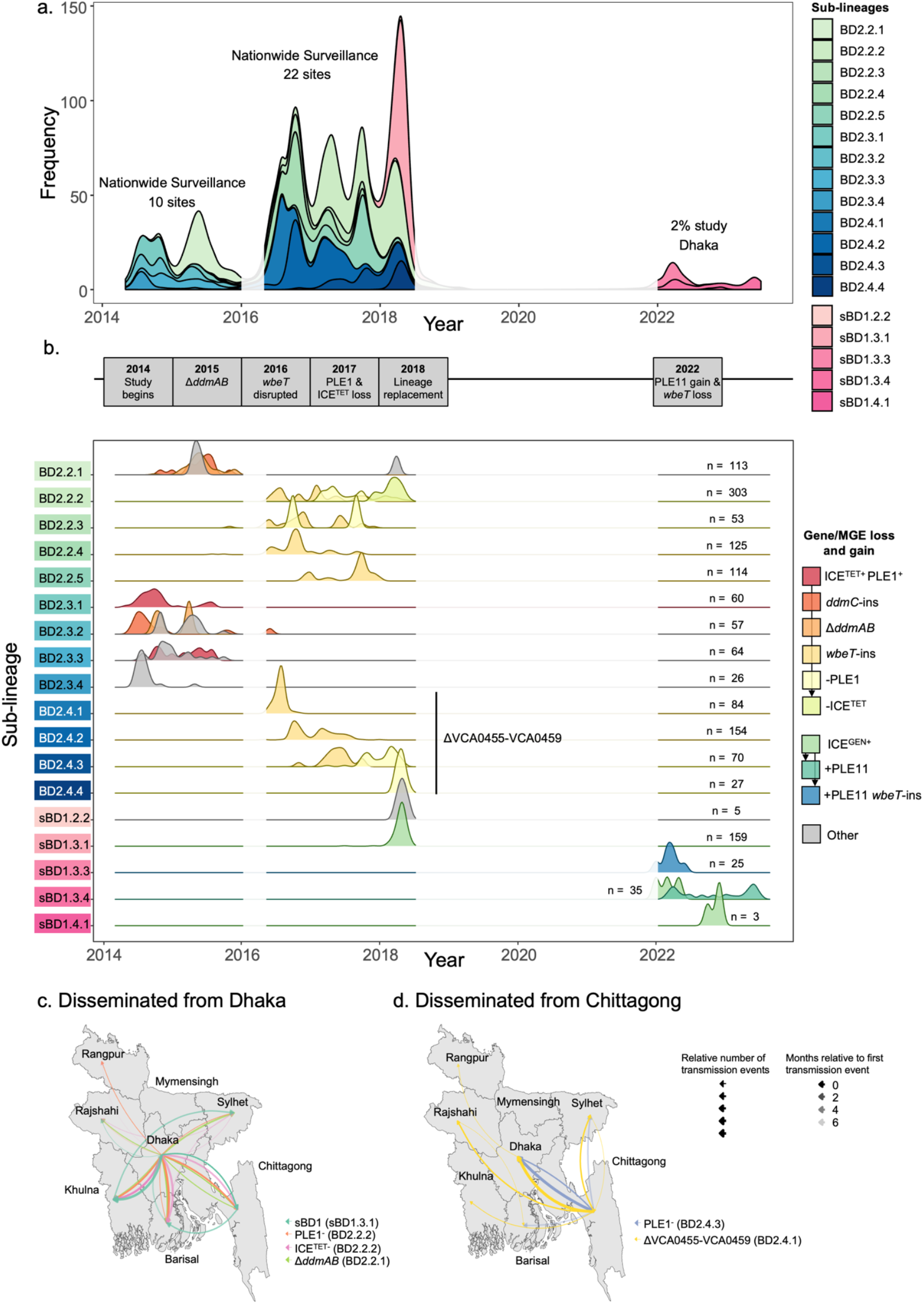
Dynamics of *V. cholerae* sub-lineages in Bangladesh and their genetic profiles over time. a. Abundance of each sub-lineage in the 2014-2018 nationwide surveillance study and 2022-2023 2% study shown over time, coloured by sub-lineage (see key). Time periods where neither the 2014-2018 surveillance study nor the 2% study were active are shaded white. b. Density plots of each sub-lineage are coloured by gene and mobile genetic element profile (see key). For each sub-lineage the total number of samples are indicated on the right. Sub-lineages within the clade lacking VCA0455-VCA0459 are indicated by a label. A timeline at the top of the figure summarises the major changes that took place each year. Time periods where neither the 2014-2018 surveillance study nor the 2% study were active are shaded white. c & d. Phylogeography of different *V. cholerae* sub-populations of interest that disseminated from Dhaka (c) and Chittagong (d). To avoid sampling bias, only samples in the 2014-2018 nationwide surveillance study were included. For each administrative division, the source of the first introduction of each sub-population to this division is indicated by an arrow. Arrows are coloured by the subpopulation of interest (see key), while their size denotes the relative number of transmission events and opacity denotes the months relative to the first transmission event (see key).

In addition to SNP variation in the core genome, *V. cholerae* genomes lost and gained individual genes and whole mobile genetic elements (Figure 1b). By the first year of the surveillance study in 2014, the complete exotoxin *hlyA* gene (VCA0219) had been lost in 81% of BD2 genomes due to a 17bp deletion and resultant frameshift mutation (Figure S4a). Furthermore, there was sequential degradation of *ddmABC*, an operon within VSP-II which triggers cell suicide during phage infection or plasmid uptake^27^. This included a frameshift mutation in *ddmC*, followed by a transposase insertion in *ddmB*, then deletion of *ddmAB* and neighbouring genes VC0493-VC0494 (Figure S4b). This was followed by disruption of *wbeT* (VC0258, Figure S4c), resulting in a phenotypic serotype switch in 2016; independent loss of anti-phage island PLE1 by two different sub-lineages in 2017, and loss of the SXT-ICE^TET^ element in 2017 (Figure 1b). Furthermore, over the course of 2016 a monophyletic clade with deletion of VCA0455*-* VCA0459 (sub-lineages BD2.4.1-BD2.4.4 in figure 1b; Figure S4d) in the superintegron became more common, comprising 51% of the BD2 isolates sequenced in 2016, but falling to 27% in 2017-2018.

Strikingly, in 2018, sBD1 sub-lineage sBD1.3.1 entirely replaced BD2 in Bangladesh. In contrast to BD2, sBD1.3.1 carried the SXT-ICE^GEN^ element, the *ctxB7* allele, a complete *hlyA* exotoxin gene, the entire *ddmABC* operon and intact *wbeT*. However, sBD1.3.1 carried a frameshift mutation in colonisation factor *acfC* (VC0841, Figure S4e) due to a nonsense mutation, and initially lacked a PLE anti-phage element. By 2022, when the 2% surveillance study in Dhaka began, three new sub-lineages had replaced sBD1.3.1, of which two were directly descended from it (sBD1.3.3 and sBD1.3.4) and another was likely introduced from India (sBD1.4.1). sBD1.4.1 was more closely related to the sub-lineages causing recent outbreaks in Pakistan, Lebanon and Malawi. In sBD1.3.4 the *wbeT* gene was disrupted by an insertion, conferring on it the Inaba serotype. Furthermore, both sBD1.3.3 and sBD1.3.4 had gained a novel PLE island, here termed PLE11, in a different genomic position to PLE1 in BD2 (Figure S5).

Next, to look for the temporal and spatial signatures in the loss and gain of gene functions in Bangladesh, we focused our analysis on genomes from the systematic 2014-2018 surveillance only to avoid sampling bias. Figure 1b shows that there are clear temporal patterns of loss: PLE1 and ICE^TET^ in sub-lineage BD2.2.2, PLE1 in BD2.4.3, *ddmAB* in BD2.2.1, and VCA0455-VCA0459 as BD2.2.4 gave rise to sub-lineage BD2.4.1. These changes were fixed in all derived sub-lineages (Figure S2). Using TreeTime, there were two patterns of geographical spread evident: sBD1.3.1, PLE1^−^ and ICE^TET-^ BD2.2.2 and Δ*ddmAB* BD2.2.1 disseminated from Dhaka (Figure 1c), while PLE1^−^ BD2.4.3 and ΔVCA0455-VCA0459 BD2.4.1. disseminated from Chittagong (Figure 1d).

In addition to the above there were also mobile genetic elements which were sporadically acquired, including plasmid pSA7G1 (Figure S6a) acquired multiple times in five BD2 sub-lineages distributed across six administrative divisions. The functional relevance of pSA7G1 is unclear. The K139 lysogenic phage, which binds to the *Vibrio cholerae* O1 antigen and carries a gene *glo* that is linked to virulence in mouse models^28,29^, was found in ten BD2 sub-lineages across seven administrative divisions, but most commonly in Chittagong (Figure S6b). Also of note, the ICP1 lytic phage was detected and sequenced from 15 *V. cholerae* samples, and like K139, was most commonly from samples taken in Chittagong (Figure S6c).

### India and Bangladesh have distinct temporal patterns of cholera

To understand the relationship between *V. cholerae* in Bangladesh and other countries falling within the Ganges basin we analysed the 794 isolate genomes collected across North India (Figure S7). Although both Bangladesh and India harboured sub-lineages falling within sBD1 and BD2, the two countries followed distinct temporal patterns of sub-lineage replacement (Figure 2a). Following co-circulation of both sBD1 and BD2 in Bangladesh and India from 2004-2011, sBD1 became predominant in India, representing 94% of samples by 2011, while BD2 became predominant in Bangladesh, representing 95% of samples by 2013. Of the 13 BD2 sub-lineages present in Bangladesh from 2013 to 2018, 12 were contained within the national boundaries of Bangladesh and not found in any other country in the world, even those bordering it or linked to the delta region. Furthermore, the PLEs that typified the sub-lineage isolates circulating in Bangladesh were rare outside the country, as was loss of *ddmABC* and complete Inaba serotype replacement (Figure S8).

**Figure 2:**
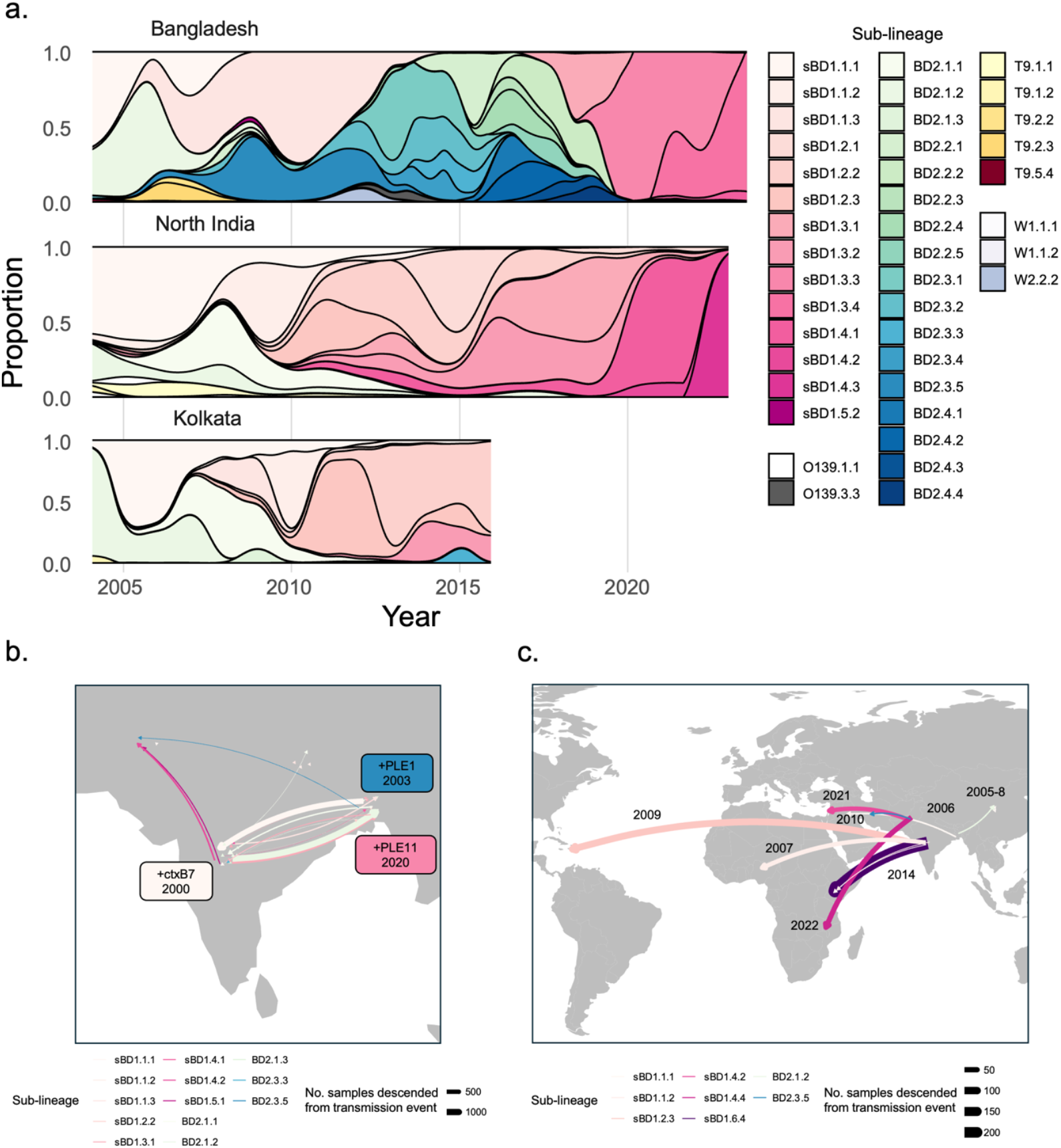
Dynamics of *V. cholerae* sub-lineages in the Ganges basin and international transmission. a. Abundance of each sub-lineage within Bangladesh, North India and Kolkata over time, including both samples collected for this study and previously published contextual samples b. Inferred transmission events and key gene/MGE gain events within South Asia. Arrows are coloured by sub-lineage, and sized according to how many samples in our global collection were descended from this transmission event. c. Inferred transmission events from South Asia to other regions. Arrows are coloured by sub-lineage. Arrows are sized according to how many samples in our global collection were descended from this transmission event, including not only the country indicated but also onwards transmission to other countries.

Conversely, the temporal patterns of *V. cholerae* seen in Kolkata aligned with North India (Figure 2a), despite Kolkata lying only 60km from the Khulna division of Bangladesh: 0/167 genomes taken in the Khulna division from 2015-2018 were in sub-lineages shared with Kolkata, and 167/167 were in sub-lineages shared with the rest of Bangladesh (Figure S9a). Similarly, all six of the samples collected in Assam (2002-2005) fell within a T9 sub-lineage present in India, despite its geographic proximity to Bangladesh (Figure S9b) and of the 12 samples collected in Nepal in 2010, 11 belonged to sub-lineages present in India at the time, and one a sub-lineage present in Bangladesh (Figure S9c).

Next, to reconstruct the history of global transmission events from the time-scaled phylogeny of global 7PET (Figure S2) we inferred the ancestral state and date of each node (Supplementary document 2). High transmission was evident between countries in South Asia, but with more transmission events between Bangladesh and India (40/408) than any other pair of countries (Figure 2b). Dissecting these events in more detail, we show that sBD1 and BD2 both separately diverged from the T9 lineage in the late 1990s. sBD1 appears to have gained the *ctxB7* allele in India, subsequently spreading to Nigeria, Kenya and ultimately to Haiti to cause the 2010 Haiti outbreak (Figure 2c). Since then, most outbreak-causing global transmission events from South Asia appear to have been seeded from the pool of *ctxB7*^+^ sBD1 evolving in India (Figure 2c). From 2003-2023 there were twice as many samples from Bangladesh compared with India in our global phylogeny, yet 32.6 times as many samples outside of South Asia were more recently descended from Indian transmission events (n = 359, 2003-2023) compared with Bangladeshi transmission events (n = 11, 2003-2023).

Despite the high number of local transmission events between India and Bangladesh, 29/40 failed to give rise to more than 10 descendent samples. In the 2010s there were seven India-to-Bangladesh transmission events evident from these data, of which six failed to persist, giving rise to fewer than 10 patient samples in Bangladesh. The subclade of sBD1.3.1 that did become eventually become predominant in Bangladesh was detected in Dhaka as early as 2015, but did not expand to displace BD2 until three years later. Conversely, although BD2 became predominant in Bangladesh from 2013-2017, from 2010 onwards there was only one observed export event, resulting in a single sample of BD2 being detected in Kolkata. Both BD2 and sBD1 gained PLEs after introduction or re-introduction to Bangladesh: BD2 was introduced to Bangladesh in approximately 2001 and gained PLE1 in Bangladesh 2.0 years later. Similarly, PLE11 was gained by sBD1 in approximately 2020, 2.0 years after sBD1 rose to >50% of samples in Bangladesh. PLE11 was not detected in any country other than Bangladesh.

### Microevolution of *Vibrio cholerae* was associated with changes in ICP1 specificity, disease severity and bacterial host mobility

Given the rapid dynamics of gene/MGE loss but comparatively low number of global transmission events originating directly from Bangladesh, we investigated how changes in genes and mobile genetic elements might affect fitness (infection potential, susceptibility to phage, and antimicrobial sensitivity) as well as the potential of different lineages to move outside of Bangladesh and the Ganges delta into India.

We first carried out a multivariate logistic regression analysis to identify whether gene or MGE losses were associated with clinical severity (Figure 3a), adjusting for the presence of other genes and MGEs. Presence of *ddmA* (p = 0.01), *wbeT* (p = 5.8×10^−4^) and ICE^TET^ (p = 0.005) were associated with reduced risk of rice water stool, and VCA0455-VCA0459 (p = 0.0001) and PLE1 (p = 0.04) with reduced risk of severe dehydration, suggesting their sequential loss may enhance disease severity. We had hypothesised that loss of these genetic elements might allow faster replication and confer a selective advantage within the gut. In the absence of ICP1, the proportion of reads in the stool metagenome occupied by *V. cholerae* rose from a median of 4.5% to 18.6% following the loss of PLE1 by *V. cholerae* carrying *ctxB1*^+^ (and therefore likely lineage BD2; Figure 3b). This further rose to 52.9% following loss of ICE^TET^ (Figure 3b). Even in the presence of ICP1 *V. cholerae* rose from 1.4% to 11.8% of the stool metagenome following ICE^TET^ loss. PLE1 and ICP1 were not detected together in any of the stool metagenomes.

**Figure 3:**
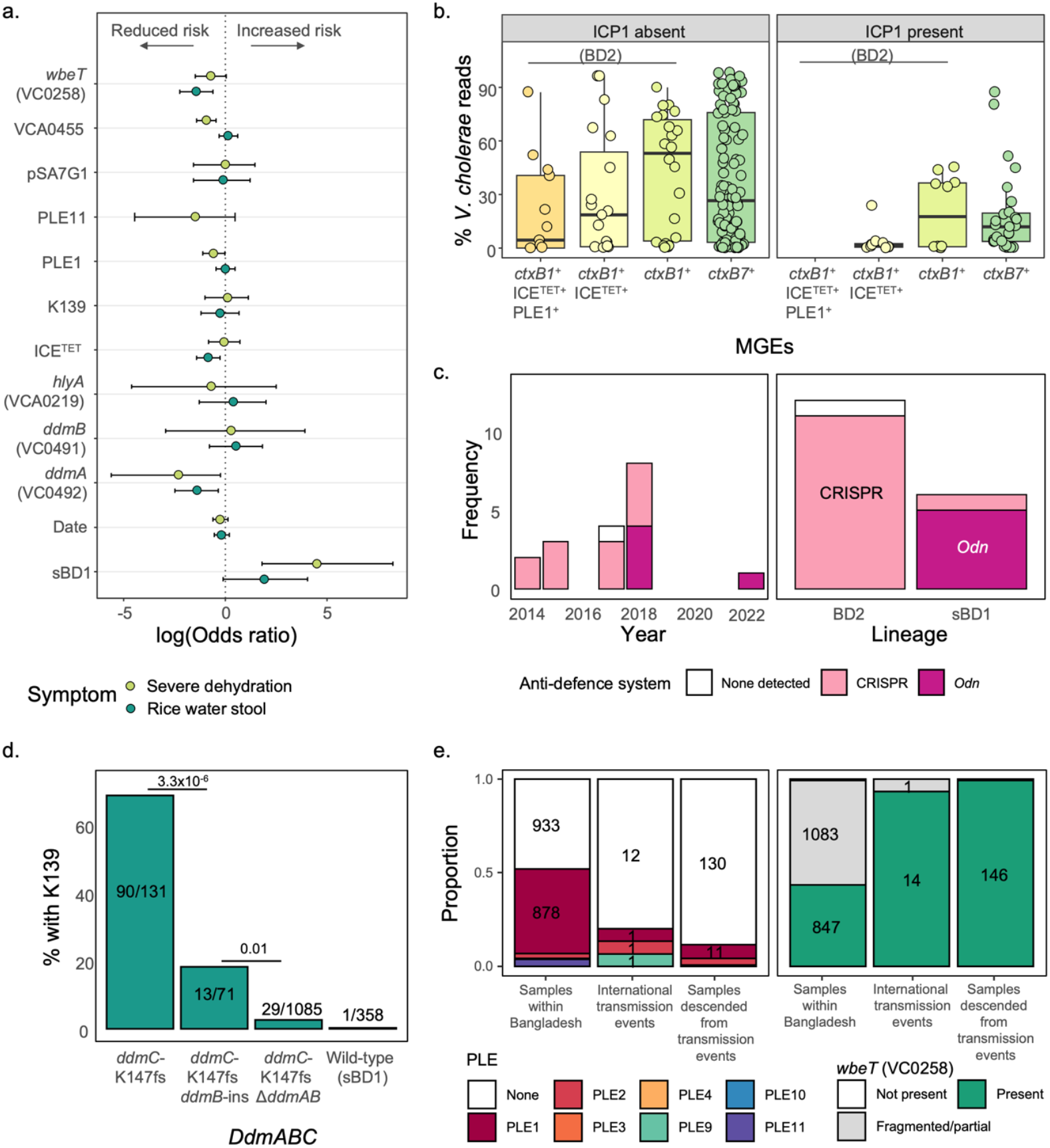
Relationships between *V. cholerae* genetic changes and phenotype. a. Log-transformed odds ratios for the association of different genes and mobile genetic elements with rice water stool and severe dehydration, adjusting for the presence/absence of other genes and mobile genetic elements, and the date, and site of collection. Error bars indicate 95% confidence intervals. b. % of reads assigned to *V. cholerae* by kraken in stool metagenomic data. Box plots indicate the median and interquartile range, with whiskers extending to the most extreme values within 1.5 x IQR. Samples were categorised by the presence of *ctxB7* and *ctxB1*, which for this time period and location delineates sBD1 and BD2, and *ctxB1*^*+*^ samples were sub-categorised by whether PLE1 and ICE^TET^ was detected. c. Frequency of ICP1 anti-defence systems detected in samples in Bangladesh, by year and by lineage of the co-sequenced *Vibrio cholerae*. d. Proportion of samples where lysogenic phage K139 were present in the genome, classified by which genes in the *ddmABC* antiviral system were present. P values are indicated for the results of a multivariate logistic regression, in which all the factors indicated in figure 3d (ICP1, pSA7G1, *ddmA* and *ddmB*, SXT-ICE, PLE, *wbeT*, VCA0455-VCA0459, *hlyA*, lineage, date and site) were tested for association with K139. e. Representation of PLE^+^ and *wbeT*^+^ *V. cholerae* among the total number of samples within Bangladesh; inferred export events, and the number of samples outside of Bangladesh descended from these export events, from 2003-2023.

Our data also showed a striking temporal link between ICP1-type and the circulating *V. cholerae*-lineage: during replacement of BD2 by sBD1 we observed a simultaneous switch in the ICP1 phage type co-sequenced alongside it, from ICP1 with a CRISPR/Cas anti-defence system (11/13 prior to April 2018) to ICP1 carrying the *odn* anti-defence gene (5/6 from April 2018 onwards) (Figure 3c). We also identified genetic changes linked to K139 phage susceptibility: surprisingly, *ddmABC* degradation was significantly associated with a fall in K139 prevalence (p = 3.3×10^−6^ and 0.01 for *ddmB* and *A* respectively, logistic regression adjusting for date, study site and presence/absence other genes, Figure 3d).

Given that PLE11 was not seen outside of Bangladesh, we then examined whether PLEs in general compromise global transmission potential. Compared to the number of PLE^+^ genomes sampled within Bangladesh, PLEs were significantly under-represented amongst transmission events to other countries from 2003-2023 (Figure 3e; p = 1.8×10^−4^, fisher test), and even more under-represented within the genomes of *V. cholerae* descended from those events (p < 2.2×10^−16^, fisher test). None (0/26) of the exported *V. cholerae* from India from 2003-2023 were PLE^+^. Similarly, Inaba serotype *V. cholerae* lacking complete *wbeT* were rarely transmitted out of Bangladesh (p = 8.4×10^−5^, fisher test).

Finally, as may be expected, phenotypic antibiotic susceptibility testing suggested that the loss of ICE^TET^ was associated with increased susceptibility to trimethoprim/sulfamethoxazole (p = 9.3×10^−8^, fisher test, Figure S10), tetracycline (p = 9×10^−4^) and doxycycline (p = 2.5×10^−5^), while sBD1 lineage *V. cholerae* carrying the ICE^GEN^ element were significantly more susceptible to doxycycline (p = 3.7×10^−22^).

## Discussion

The Sundarbans and highly populated areas within the Ganges delta region of Bangladesh, bordering the Bay of Bengal, are considered by many to be the heartland of epidemic cholera^1^. This notion is reinforced by genomic studies showing that a discrete clone of *V. cholerae* named 7PET radiated from this region in multiple epidemic waves to cause the ongoing seventh pandemic^12^. It is 7PET that is responsible for the current increase in cholera globally, and the reason that the WHO has declared cholera resurgence as a grade 3 emergency^30^. Whilst we are aware of the symptoms of global spread, with 667,000 cholera cases and 5/6 WHO regions reporting outbreaks in 2023^31^, we know little about the evolutionary dynamics of *V. cholerae* at its global source. Here we used genomics to understand the dynamics of *V. cholerae* across the greater Ganges region, including both the delta regions within Bangladesh and India as well as the upper Ganges Basin covering eight North Indian states. We followed the evolution of sBD1 and BD2, which were the dominant circulating 7PET lineages in this region in this time.

Our results show that despite the Ganges delta and basin spanning Bangladesh and India, cholera evolution follows national borders rather than its hydrological features and flow. Both Kolkata and Assam harboured cholera sub-lineages present in North India but not Bangladesh, while in Bangladesh the evolution of BD2 occurred almost in isolation from the rest of the world. Furthermore, many of the genetic changes in BD2 appeared to spread nationwide from Dhaka, where population density is greatest. Although there may be bias, even within the context of a systematic surveillance study, together with following national borders this adds to the overwhelming evidence that 7PET cholera transmission is primarily mediated by short-cycle human-human transmission.

*V. cholerae* evolution in Bangladesh was characterised by rapid changes in genes and mobile genetic elements, particularly those relating to phage defence. Sub-lineages harbouring these changes rapidly became dominant, suggesting that *V. cholerae* evolution is driven by strong selective pressures. This is illustrated by the almost simultaneous switch in *V. cholerae* phage defence systems and ICP1 anti-defence systems in 2018. This strongly suggests that in Bangladesh ICP1 lytic phage plays a major role in cholera evolution. It is possible that once ICP1 co-evolved to overcome phage defences in BD2, retaining these systems no longer presented an evolutionary advantage, resulting in their sequential loss. PLE1 was independently lost at multiple points in BD2’s evolution, signifying that its maintenance presented a strong burden. A similar pattern was observed for the anti-phage *ddmABC* operon: once a frameshift mutation in *ddmC* rendered the operon ineffective^32^, the rest of the operon was deleted. Having lost its defences, the BD2 lineage was then rapidly replaced by sBD1, harbouring an ICE^GEN^ element containing an anti-phage BREX system^10^ that was likely initially effective against local ICP1. However, ICP1 then rapidly switched from a CRISPR to an *Odn* anti-defence system. Finally, sBD1 overcomes this through acquisition of novel anti-phage island PLE11, which prevents ICP1 propagation by disrupting tail assembly, therefore acting as a strong evolutionary driver for ICP1 evolution^33^. Further, our data reveals the carriage of these phage defence systems were associated with lower disease severity and *V. cholerae* load, suggesting that maintenance of these systems has a significant cost to pathogenicity. The loss of ICE^TET^, which confers tetracycline resistance, indicates that antibiotic treatment on the other hand does not exert as strong an evolutionary pressure as ICP1.

Interestingly, while PLE anti-phage islands were rare outside of Bangladesh, both BD2 and sBD1 gained a PLE approximately two years after becoming prevalent in Bangladesh. Furthermore, PLEs were under-represented amongst transmission events out of Bangladesh, suggesting they may only be advantageous within the Ganges delta, and compromise long-range transmission within and outside of South Asia. This may partially explain why the inferred source of most global transmission events appears to be India rather than Bangladesh, refining the previous notion that the Ganges delta holds the global diversity of all epidemic 7PET *V. cholerae* sub-lineages. Rather, surrounding countries including India are the launchpads for global transmission.

It is currently unclear why PLEs appear to be largely confined to Bangladesh, as ICP1 has also been found in India, Yemen and the Democratic Republic of Congo^5,34,35^. It is possible that the year-round endemicity of *V. cholerae*, as well as high salinity drinking water which creates conditions favourable for ICP1 infection^36^, could allow maintenance of greater ICP1 population diversity in Bangladesh than other countries. Interestingly, we found that both K139 and ICP1 phages appeared to be most common in Chittagong, a division with both high *V. cholerae* prevalence^37^ and increasing saltwater inundation of drinking water^38^. Our hypothesis is that Bangladesh is one of the few places with a high enough human population density and cholera prevalence, combined with high exposure rates due to a lack of preventative WASH infrastructure, to effectively transmit within this population while maintaining bulky anti-phage defence systems. Furthermore, we found that Inaba serotype *V. cholerae* rarely transmitted outside of Bangladesh, suggesting that while serotype-switching may confer immune evasion in populations with high Ogawa-specific seroprevalence^39,40^, it could also present a fitness cost in immune-naïve populations.

In summary, we found that *V. cholerae* evolution in Bangladesh was unique in its rapid gain and loss of mobile genetic elements. This followed relatively predictable patterns, gaining PLEs when becoming established in Bangladesh, and losing them over time, likely driven by the trade-off between maintaining bulky anti-phage defences against ICP1 and pathogenicity: classical gene-for-gene theory but here on a regional scale^23,41,42^. Furthermore, within Bangladesh *V. cholerae* appeared to radiate from areas of high population density such as Dhaka and Chittagong. Real-time, longitudinal genomic surveillance of *V. cholerae* in these regions could be utilised as early-warning system for the entire country, identifying emerging sub-lineages harbouring changes predicted to increase risk of severe disease or antibiotic resistance. This could allow rapid mobilisation of intervention strategies, such as vaccination or WASH, to prevent dissemination of high-risk sub-lineages. We also found that MGEs such as PLEs could compromise the global transmission potential and export of *V. cholerae* from Bangladesh into the upper Ganges region. Thus despite the high prevalence and diversity of cholera in Bangladesh, the Ganges delta may not represent the modern global source of cholera, but rather India and the Ganges basin as a whole. From our data it is clear that focusing cholera surveillance on outbreaks such as in Africa, the Middle East, and Haiti will be insufficient to end the seventh cholera pandemic. Global annual cholera surveillance, akin to the surveillance and response system currently in place for influenza, will be necessary to effectively target sources of transmission using WASH interventions and the limited oral cholera vaccine stockpile.

## Materials and methods

### Bangladesh sample collection and sequencing

Study procedures for the 2014-2018 surveillance study are outlined in detail in Khan et al. 2020^43^, and were approved by the Research Review Committee and Ethical Review Committee of icddr,b. Briefly, sentinel surveillance was carried out at ten sites from 2014-2016, interrupted from January to May 2016 due to a gap in funding, followed by extension to 22 sites in 21 districts from May 2016 onwards. Informed written consent was obtained from all adult participants, or from legal guardians for children younger than 18 years old. A stool sample was collected for detection of *Vibrio cholerae* O1/O139. At the ten sites where surveillance was established in 2014, samples were also tested for enterotoxigenic *Escherichia coli* (ETEC), *Salmonella* and *Shigella* species. Among 26,221 acute watery diarrhoea patients enrolled, 6.2% (n = 1604) cases were *V. cholerae* O1, of which 1526 were whole genome sequenced. Similarly, from the 2% icddr,b Dhaka Hospital Surveillance - where every 50th patient visiting the icddr,b Hospital in Dhaka was enrolled and tested for enteric pathogens - 63 *Vibrio cholerae* samples from 2022-23 were also sequenced.

From the sentinel sites, stool samples were transported in Cary-Blair media to icddr,b within 15 days of sample collection^43^. Samples were streaked directly onto taurocholate-tellurite gelatin agar (TTGA), and also enriched in alkaline peptone water (APW, pH 8.6) for 18 hours and plated on TTGA. Samples were incubated overnight at 37°C. Suspected *V. cholerae* colonies were serotyped using O1 Ogawa-, O1 Inaba- and O139-specific antibodies. A subsample (every 5^th^ *V. cholerae* positive culture) underwent antibiotic susceptibility testing (doxycycline n = 311, tetracycline n = 195, erythromycin n = 366, azithromycin n = 350, ciprofloxacin n = 350, trimethoprim/sulfamethoxazole n = 195, nalidixic acid n = 140) using commercially available antibiotic discs (Oxoid, Basingstoke, United Kingdom) following the guidelines of the Clinical and Laboratory Standards Institute^44^. In 2017, testing for ampicillin (n = 187), ceftriaxone (n = 171) and cefixime (n = 171) was introduced. *Escherichia coli* American Type Culture Collection 25922 susceptible to all antimicrobials was used as a control strain.

Genomic DNA was extracted from 5ml cultures of *V. cholerae* incubated overnight at 37°C in Luria-Bertani (LB) media, using the Wizard Genomic DNA Kit (Promega, Madison, WI, USA). DNA integrity was confirmed by agarose gel electrophoresis, and purity using a NanoDrop™ 2000 Spectrophotometer (Thermo Fisher Scientific, USA). 150bp paired-end sequencing was carried out at the Wellcome Sanger Institute using the Illumina Hiseq2500 platform (Illumina, San Diego, CA, USA). Fastq files were trimmed using fastp v0.23.4, moving a sliding window from the 5’ and 3’ end of the reads and trimming bases with a mean quality below 20. Read quality was then verified using FastQC v0.11.8 and MultiQC v1.8.

### North India sample collection and sequencing

This study was approved by the Institute Ethics Committee, PGIMER-Chandigarh. Stool and water samples were collected during an outbreak investigation in affected areas within Chandigarh and neighbouring states in North India. Additionally, water samples were obtained from freshwater sites, including rivers and ponds. Stool samples and water samples were transported in Cary-Blair media and under a cold chain, respectively, to PGIMER, Chandigarh, for processing.

Water samples were filtered using 0.22 μm nitrocellulose acetate filters, followed by vortexing in phosphate-buffered saline (PBS) to release adherent cells. Both filtered water samples (100 μL aliquots) and stool samples were enriched in alkaline peptone water (APW) at 37°C for 6–8 hours. This was followed by subculturing the enriched samples onto blood agar and thiosulfate-citrate-bile salts-sucrose (TCBS) agar for further incubation at 37°C for 18–24 hours. Collected samples were also tested for other enteric pathogens, such as *Shigella* and *Salmonella*. Colonies resembling *Vibrio cholerae* or other enteric pathogens were identified using MALDI-TOF. Genomic DNA was extracted using as above. High-throughput genome sequencing was carried out on the Illumina platform to generate 150 bp paired-end reads, and quality control of the sequencing data was performed as described above.

### Phylogenetic analysis

We contextualised these resulting genomes within a global collection of 7PET genomic sequences (supplementary document 1). Reads were mapped against *V. cholerae* N16961, using snippy v4.6^45^ to create a pseudo-genome alignment. Snp-sites v2.5.1^46^ was used to create a SNP-only alignment, and snp-dists v0.7^47^ to calculate pairwise distances between genomes. Kraken v1.1.1^48^ was used to find the proportion of reads attributed to *V. cholerae* and ICP1. Samples with a SNP distance >400 from reference N16961 or <90% reads attributed to *V. cholerae* were excluded. 7PET samples were hierarchical clustered on the basis of their pseudo-genome alignment using rHierBAPS v1.1.3^49^, setting the maximum number of populations to thirteen and depth to three. First-level clusters were considered to represent major 7PET lineages such as sBD1 and BD2. Lineages were further divided into sub-lineages on the basis of second and third-level clusters. Maximum likelihood phylogenetic trees for a) Global 7PET, b) sBD1 within the systematic 2014-2018 surveillance study, and b) BD2 within the systematic 2014-2018 surveillance study, were created using IQ-tree v1.6.12 using the HKY+F+I substitution model^50^. TreeTime v0.7.4^51^ was used to re-root trees and create a maximum-likelihood time-scaled phylogeny. The discrete ancestral state of each node, including location, sub-lineage and the presence of key genes and mobile genetic elements, was estimated using TreeTime ‘mugration’. Transitions between different ancestral locations in the global 7PET tree were used to estimate transmission between different countries, and transitions between locations in the sBD1 and BD2 systematic 2014-2018 surveillance study trees to infer transmission events between different divisions within Bangladesh. Phylogenetic trees were visualized using the R packages ape v5.8^52^ and ggtree v3.12.0^53^.

### *De novo* Genome Assembly and Analysis

*Vibrio cholerae* genome assembly was carried out using the WSI Pathogens assembly pipeline^54^. Briefly, sequences were assembled into contigs using Velvet v1.2.10^55^ followed by assembly improvement using SSPACE^56^ and GapFiller^57^. To assess assembly, quality-controlled reads (see above) were aligned to the final assembly using SMALT (https://sourceforge.net/projects/smalt/). Assemblies were annotated using PROKKA v1.5^58^.

### Virulence genes, antibiotic resistance and mobile genetic elements

ARIBA v2.14.6^59^ was used to detect the presence of complete reading frames for antibiotic resistance genes (against the CARD database), virulence factors (against the VFDB Virulence Factor database), *ctxB* types, all genes present in the N16961 reference genome, and ICP1 anti-defence genes (reference genome MW794190.1 for *csy1-4, cas1* and *cas3* genes, MW794192.1 for the *odn* gene). Mash v2.1.1^60^ was used to screen for sample read sets contained within mobile genetic element sequences, including PLE1 (KC152960.1), PLE2 (KC152961.1), PLEs 3-10^32^, PLE11 (identified from *de novo* genome assemblies using Panaroo v1.3.4^61^), SXT-ICE^TET^ (MK165649.1), SXT-ICE^GEN^/ICEVchInd5 (KY382507.1), and plasmids pCNRVC190243^5^ (OW443149.1) and pSAG71 (CP053818.1). A mobile genetic element was considered present if a sample shared 700/1000 hashes. Samples co-sequenced with ICP1 were identified based on >0.1% of reads mapping to ICP1 using kraken v1.1.1. Association of key genes and MGEs with rice water stool, severe dehydration and K139 prevalence was assessed using a multivariate logistic regression model, implemented in R adjusting for presence of all other key genes/MGEs, time and sampling site.

### Secondary metagenomic analysis

Publicly available metagenomic data from participants infected with or exposed to *Vibrio cholerae* in Bangladesh (BioProjects PRJNA608678 and PRJNA976726)^62,63^ were downloaded (n = 426 metagenomes). Kraken v1.1.1 was used to quantify the percentage of reads mapped to *Vibrio cholerae* and ICP1. As above, ARIBA v2.14.6 was used to detect the presence of individual genes and *ctxB* alleles, and Mash v2.1.1 to detect MGEs.

## Data availability

The read data generated in this study have been deposited in the ENA database under the accession IDs indicated in supplementary document 1.

## Acknowledgment

This work was supported by the Bill and Melinda Gates Foundation. We are thankful to officials from the Government of Bangladesh, icddr,b physicians, and the participants who took part in interviews that provided support to this analysis. icddr,b is grateful to the governments of Bangladesh and Canada, for providing core/unrestricted support. PGIMER-Chandigarh (India) acknowledge the help and support provided by local health authorities in investigating and managing the outbreaks of cholera. Sequencing was supported by the Wellcome Sanger Institute. A.B., A.T.B. and N.R.T. were supported by Wellcome funding to the Sanger Institute (grants 108413/A/15/D and 206194). For the purpose of open access, the authors have applied a CC-BY public copyright licence to any Author-Accepted Manuscript version arising from this submission.

## Competing interests

The authors declare no competing interests.

## Contributions

A.B., M.H.A., A.T.B, N.R.T. and F.Q. conceived the work and designed the study. M.H.A., T.I., S.I.A.R, M.A., Y.A.B., T.R.B., and A.I.K. were involved in acquisition of samples and data in Bangladesh, and N.S., C.T. and N.T. in North India. A.T.B. compiled the global collection of cholera genomes. A.B. performed data analysis and visualisation. A.B., M.H.A., N.S. and C.T. wrote the manuscript. All authors contributed to interpretation of the data and results and revised the manuscript. All authors approved the final version of the manuscript.

## Notes

### Competing Interest Statement

The authors have declared no competing interest.

### Funding Statement

This study was funded by Wellcome funding to the Sanger Institute (grants 108413/A/15/D and 206194), the Bill and Melinda Gates Foundation, and the governments of Bangladesh and Canada.

### Author Declarations

The Research Review Committee and Ethical Review Committee of the International Centre for Diarrhoeal Disease Research, Bangladesh, and Institute Ethics Committee of the Postgraduate Institute of Medical Education and Research, Chandigarh, gave ethical approval for this work.

## References

1. Siddique, A. K. & Cash, R. Cholera Outbreaks in the Classical Biotype Era. in Cholera Outbreaks (eds. G. Balakrish Nair & Yoshifumi Takeda) (2014).

2. Ryan, E. T. The cholera pandemic, still with us after half a century: Time to rethink. PLoS Neglected Tropical Diseases vol. 5 Preprint at 10.1371/journal.pntd.0001003 (2011).

3. Chin, C.-S. et al. The Origin of the Haitian Cholera Outbreak Strain. New England Journal of Medicine 364, 33–42 (2011).

4. Weill, F. X. et al. Genomic insights into the 2016–2017 cholera epidemic in Yemen. Nature 565, 230–233 (2019).

5. Lassalle, F. et al. Genomic epidemiology reveals multidrug resistant plasmid spread between Vibrio cholerae lineages in Yemen. Nat Microbiol 8, 1787–1798 (2023).

6. Abou Fayad, A. et al. An unusual two-strain cholera outbreak in Lebanon, 2022-2023: a genomic epidemiology study. Nat Commun 15, (2024).

7. United Nations International Children’s emergency funds [UNICEF]. Syria Cholera Response Situation Report for 04 October 2022 - Syrian Arab Republic. https://www.unicef.org/syria/reports/cholera-situation-report-4-october-2022 (2022).

8. Chaguza, C. et al. Genomic insights into the 2022–2023 Vibrio cholerae outbreak in Malawi. Nat Commun 15, (2024).

9. Monir, M. M. et al. Genomic attributes of Vibrio cholerae O1 responsible for 2022 massive cholera outbreak in Bangladesh. Nat Commun 14, 1–8 (2023).

10. Chun, J. et al. Comparative genomics reveals mechanism for short-term and long-term clonal transitions in pandemic Vibrio cholerae. PNAS (2009).

11. Hu, D. et al. Origins of the current seventh cholera pandemic. Proc Natl Acad Sci U S A 113, E7730–E7739 (2016).

12. Mutreja, A. et al. Evidence for several waves of global transmission in the seventh cholera pandemic. Nature 477, 462–465 (2011).

13. Weill, F. X. et al. Genomic history of the seventh pandemic of cholera in Africa. Science (1979) 358, 785–789 (2017).

14. Domman, D. et al. Integrated view of Vibrio cholerae in the Americas. Science (1979) 793, 789–793 (2017).

15. Oprea, M. et al. The seventh pandemic of cholera in Europe revisited by microbial genomics. Nat Commun 11, (2020).

16. Weill, F. X. et al. Genomic history of the seventh pandemic of cholera in Africa. Science (1979)358, 785–789 (2017).

17. Monir, M. M. et al. Genomic Characteristics of Recently Recognized Vibrio cholerae El Tor Lineages Associated with Cholera in Bangladesh, 1991 to 2017. Microbiol Spectr 1–13 (2022).

18. Morita, D. et al. Whole-Genome Analysis of Clinical Vibrio cholerae O1 in Kolkata, India, and Dhaka, Bangladesh, Reveals Two Lineages of Circulating Strains, Indicating Variation in Genomic Attributes. mBio 11, 1–9 (2020).

19. Baddam, R. et al. Genome dynamics of Vibrio cholerae isolates linked to seasonal outbreaks of cholera in Dhaka, Bangladesh. mBio 11, 1–14 (2020).

20. Imamura, D. et al. Comparative genome analysis of VSP-II and SNPs reveals heterogenic variation in contemporary strains of Vibrio cholerae O1 isolated from cholera patients in Kolkata, India. PLoS Negl Trop Dis 11, 1–14 (2017).

21. Taylor-Brown, A. et al. Genomic epidemiology of Vibrio cholerae during a mass vaccination campaign of displaced communities in Bangladesh. Nat Commun 14, 1–11 (2023).

22. Krin, E. et al. Systematic transcriptome analysis allows the identification of new type I and type II Toxin/Antitoxin systems located in the superintegron of Vibrio cholerae. Res Microbiol 174, (2023).

23. LeGault, K. N. et al. Temporal shifts in antibiotic resistance elements govern phage-pathogen conflicts. Science (1979) 373, 1–29 (2021).

24. Seed, K. D., Lazinski, D. W., Calderwood, S. B. & Camilli, A. A bacteriophage encodes its own CRISPR/Cas adaptive response to evade host innate immunity. Nature 494, 489–491 (2013).

25. Domman, D. et al. Integrated view of Vibrio cholerae in the Americas. Science (1979) 793, 789–793 (2017).

26. Ramamurthy, T. et al. Vibrio cholerae O139 genomes provide a clue to why it may have failed to usher in the eighth cholera pandemic. Nat Commun 13, 1–10 (2022).

27. Jaskólska, M., Adams, D. W. & Blokesch, M. Two defence systems eliminate plasmids from seventh pandemic Vibrio cholerae. Nature 604, 323–329 (2022).

28. Nesper, J., Blaß, J., Fountoulakis, M. & Reidl, J. Characterization of the major control region of Vibrio cholerae bacteriophage K139: Immunity, exclusion, and integration. J Bacteriol 181, 2902–2913 (1999).

29. Reidl, J. & Mekalanos, J. J. Characterization of Vibrio cholerae bacteriophage K139 and use of a novel mini-transposon to identify a phage-encoded virulence factor. Mol Microbiol 18, 685– 701 (1995).

30. The Lancet. Cholera: a pandemic ignored. The Lancet 404, 1724–1725 (2024).

31. World Health Organisation. Multi-Country Outbreak of Cholera, External Situation Report #10. https://idsp.nic.in/index4.php?lang=1&level=0&linkid=406&lid=3689. https://idsp.nic.in/index4.php?lang=1&level=0&linkid=406&lid=36 (2024).

32. O’Hara, B. J., Alam, M. & Ng, W. L. The Vibrio cholerae Seventh Pandemic Islands act in tandem to defend against a circulating phage. PLoS Genet 18, (2022).

33. Mathur, Y. et al. Capturing dynamic phage-pathogen coevolution by clinical surveillance. Preprint at 10.1101/2025.01.29.635557 (2025).

34. Boyd, C. M. et al. Bacteriophage ICP1: A Persistent Predator of Vibrio cholerae. 23, 28 (2024).

35. Alam, M. et al. Emergence and Evolutionary Response of Vibrio cholerae to Novel Bacteriophage, Democratic Republic of the Congo. Emerg Infect Dis 2482–2490 (2022).

36. Silva-Valenzuela, C. A. & Camilli, A. Niche adaptation limits bacteriophage predation of Vibrio cholerae in a nutrient-poor aquatic environment. Proc Natl Acad Sci U S A 116, 1627–1632 (2019).

37. Khan, A. I. et al. Epidemiology of cholera in Bangladesh: Findings from nationwide hospital-based surveillance, 2014-2018. Clinical Infectious Diseases 71, 1635–1642 (2020).

38. Ashrafuzzaman, M., Artemi, C., Duarte Santos, F. & Schmidt, L. Current and Future Salinity Intrusion in the South-Western Coastal Region of Bangladesh. Span. J. Soil Sci. (2022).

39. Ali, M., Emch, M., Park, J. K., Yunus, M. & Clemens, J. Natural cholera infection-derived immunity in an endemic setting. Journal of Infectious Diseases 204, 912–918 (2011).

40. Khan, A. I. et al. Comparison of clinical features and immunological parameters of patients with dehydrating diarrhoea infected with Inaba or Ogawa serotypes of Vibrio cholerae O1. Scand J Infect Dis 42, 48–56 (2010).

41. Flor, H. H. Current Status of the Gene-For-Gene Concept. Annu Rev Phytopathol 9, 275–296 (1971).

42. Angermeyer, A. et al. Evolutionary Sweeps of Subviral Parasites and Their Phage Host Bring Unique Parasite Variants and Disappearance of a Phage CRISPR-Cas System. mBio 13, e0308821 (2021).

43. Khan, A. I. et al. Epidemiology of cholera in bangladesh: Findings from nationwide hospital-based surveillance, 2014-2018. Clinical Infectious Diseases 71, 1635–1642 (2020).

44. Clinical and Laboratory Standards Institute. Performance Standards for Antimicrobial Susceptibility Testing. (2018).

45. Seemann, T. Snippy: rapid haploid variant calling and core SNP phylogeny. Preprint at github.com/tseemann/snippy (2015).

46. Page, A. J. et al. SNP-sites: rapid efficient extraction of SNPs from multi-FASTA alignments. Microb Genom 2, e000056 (2016).

47. Seemann, T. Pairwise SNP distance matrix from a FASTA sequence alignment. Preprint at https://github.com/tseemann/snp-dists (2018).

48. Wood, D. E. & Salzberg, S. L. Kraken: ultrafast metagenomic sequence classification using exact alignments. Genome Biol (2014).

49. Tonkin-Hill, G., Lees, J. A., Bentley, S. D., Frost, S. D. W. & Corander, J. RhierBAPs: An R implementation of the population clustering algorithm hierbaps. Wellcome Open Res 3, (2018).

50. Nguyen, L. T., Schmidt, H. A., Von Haeseler, A. & Minh, B. Q. IQ-TREE: A fast and effective stochastic algorithm for estimating maximum-likelihood phylogenies. Mol Biol Evol 32, 268– 274 (2015).

51. Sagulenko, P., Puller, V. & Neher, R. A. TreeTime: Maximum-likelihood phylodynamic analysis. Virus Evol 4, (2018).

52. Paradis Emmanuel. ape: Analyses of Phylogenetics and Evolution. Preprint at 10.32614/CRAN.package.ape (2024).

53. Yu, G., Smith, D. K., Zhu, H., Guan, Y. & Lam, T. T. Y. Ggtree: an R Package for Visualization and Annotation of Phylogenetic Trees With Their Covariates and Other Associated Data. Methods Ecol Evol 8, 28–36 (2017).

54. Wellcome Sanger Institute Pathogen Informatics. vr-codebase. https://github.com/sanger-pathogens/vr-codebase.

55. Zerbino, D. R. & Birney, E. Velvet: Algorithms for de novo short read assembly using de Bruijn graphs. Genome Res 18, 821–829 (2008).

56. Boetzer, M., Henkel, C. V., Jansen, H. J., Butler, D. & Pirovano, W. Scaffolding pre-assembled contigs using SSPACE. Bioinformatics 27, 578–579 (2011).

57. Nadalin, F., Vezzi, F. & Policriti, A. GapFiller: A de novo assembly approach to fill the gap within paired reads. BMC Bioinformatics 13, (2012).

58. Seemann, T. Prokka: Rapid prokaryotic genome annotation. Bioinformatics 30, 2068–2069 (2014).

59. Hunt, M. et al. ARIBA: Rapid antimicrobial resistance genotyping directly from sequencing reads. Microb Genom 3, 1–11 (2017).

60. Ondov, B. D. et al. Mash: Fast genome and metagenome distance estimation using MinHash. Genome Biol 17, (2016).

61. Tonkin-Hill, G. et al. Producing polished prokaryotic pangenomes with the Panaroo pipeline. Genome Biol 21, (2020).

62. Madi, N. et al. Phage predation, disease severity, and pathogen genetic diversity in cholera patients. Science (1979) 384, (2024).

63. Levade, I. et al. Predicting Vibrio cholerae Infection and Disease Severity Using Metagenomics in a Prospective Cohort Study. Journal of Infectious Diseases 223, 342–351 (2021).

